# Performance of Road-Traffic-Based Exposure Proxies Against Personal PM_2.5_ Measurements in Three Sub-Saharan African Countries

**DOI:** 10.64898/2026.03.13.26348337

**Authors:** Handsome Bongani Nyoni, Terence Darlington Mushore, Laura Munthali, Sibusisiwe Audrey Makhanya, Laurine Chikoko, Stanley Luchters, Matthew F Chersich, Fortunate Machingura, Liberty Makacha, Benjamin Barratt, Hiten D Mistry, Marie Laure Volvert, Peter von Dadelszen, Anna Roca, Umberto D’alessandro, Marleen Temmeran, Esperança Sevene, Tamara Govindasamy, Prestige Tatenda Makanga, The PRECISE Network, The HE^2^AT Centre

**Affiliations:** Place Alert Labs, Surveying and Geomatics Department, Faculty of the Built Environment, Art and Design, Midlands State University, Gweru, Zimbabwe; Department of Architecture, Planning and Geomatics, University of Cape Town, Cape Town 7700, South Africa; Climate Environment and Health Department, Center for Sexual Health and HIV AIDS Research (CeSHHAR), Harare, Zimbabwe; IBM Research Africa, South Africa; Department of International Public Health, Liverpool School of Tropical Medicine (LSTM), Liverpool, UK; Department of Women and Children’s Health, School of Life Course and Population Sciences, Faculty of Life Sciences & Medicine, King’s College London, United Kingdom; Environmental Research Group, MRC Centre for Environment and Health, Michael Uren Biomedical Engineering Hub, White City Campus, Imperial College London, United Kingdom; Department of Obstetrics and Gynaecology, University of British Columbia, Vancouver, British Columbia, Canada; Department of Public Health and Primary Care, Ghent University, Belgium; Department of Space Science and Applied Physics, Faculty of Sciences, University of Zimbabwe; Malawi Liverpool Wellcome Programme, Public Health Group, Blantyre, Malawi; Medical Research Council Unit The Gambia at the London School of Hygiene and Tropical Medicine, Fajara, The Gambia; Centre of Excellence Women and Child Health, Aga Khan University, Nairobi, Kenya; Eduardo Mondlane University, Faculty of Medicine and Centro de Investigação em Saúde da Manhiça (CISM), Mozambique; Department of Public Health and Epidemiology, College of Life Sciences, University of Leicester; Trinity College, Dublin, Ireland; Wits Planetary Health Research, Wits Health Consortium, South Africa

## Abstract

**Introduction:** Particulate Matter (PM_2.5_) exposure contributes to the global disease burden, yet its monitoring remains sparse and uneven and is limited in many limited ground monitoring network settings. Road-traffic proxy indicators can provide indirect estimates of PM_2.5_ where measurements are limited but require context-specific validation. We evaluated three PM_2.5_ road-traffic related proxies:(I) population-Weighted Road Network Density (WRND), (ii) Euclidean (straight line) distance from highways (EH), and (iii) Euclidean distance from main roads (EM).

**Methods:** We validated proxies using high-resolution outdoor filtered PM_2.5_ personal exposure measurements collected over 1 year from 343 postpartum participants in The Gambia, Kenya, and Mozambique. Village-level spatial patterns for the PM_2.5_-proxy relationship were mapped using 5 km hexagonal aggregated tessellations. Proxy-PM_2.5_ associations were assessed using Spearman correlation, and predictive utility was tested using country-specific and global Random Forest (RF) models (3-fold cross-validation), reporting R^2^, RMSE, and feature importance

**Results:** Spatial mapping showed heterogeneous proxy–PM_2.5_ relationships across and within sites, with elevated PM_2.5_ occurring in both low- and high-proxy contests. WRND–PM_2.5_ correlations were weak overall and statistically significant only in Mozambique (r = 0.351; *p = 0.005*), with non-significant associations in Kenya (r = −0.041; *p = 0.673*) and The Gambia (r = −0.020; *p = 0.909*). EH–PM_2.5_ correlations were positive in The Gambia (r = 0.335; *p = 0.053*) and Mozambique (r = 0.292; *p = 0.020*) but negative and significant in Kenya (r = −0.224; *p = 0.018*).Single-variable RF models performed poorly across all countries (R^2^ < 0.45) and the Global model (R^2^=0.42). Combining proxies improved performance in Kenya (R^2^=0.52; RMSE=31.7µg/m^3^) and Mozambique (R^2^=0.60; RMSE=8.9 µg/m^3^), Global R^2^=0.46; RMSE=29.1 µg/m^3^), although in The Gambia, the combined model (R^2^=0.53; RMSE=37.6 µg/m^3^) did not exceed the best single-proxy model.

**Conclusion:** Road-network proxies provide context-dependent signals of personal PM_2.5_ exposure, and predictive performance is strengthened when proxies are combined in a hybrid model.

## 1 Introduction

Exposure to ambient and household air pollution, including fine Particulate Matter with an aerodynamic diameter ≤2.5 micrometers (PM_2.5_), remains a significant global public health concern in the 21st century. PM_2.5_ exposure contributes to an estimated 6.7 million premature deaths annually (1,2). These deaths are driven by cardiopulmonary pathways, including asthma, hypertensive disorders of pregnancy such as pre-eclampsia, cardiovascular disease, and a range of acute and chronic respiratory outcomes (3–5). The substantial health burden is partly attributable to PM_2.5_ size and chemical composition, which facilitate deep penetration into the respiratory tract, translocation across biological barriers, and systemic inflammation that affects multiple organ systems (3,4,6).Emerging evidence also links prenatal PM_2.5_ exposure to preterm birth, low birth weight, and impaired neurodevelopment among children (7,8). While a growing body of recent works suggests associations with poor mental well-being and general health (9–11). These impacts are amplified in sub-Saharan Africa (SSA) by rapid urbanization, increased motorization, unpaved-road dust, climate change, primary production, and landscape fire smoke, which collectively intensify both primary emissions and secondary particle formation (12,13). Effective management and mitigation of PM_2.5_ exposure depend on reliable air quality monitoring stations capable of capturing spatiotemporal variability (14). However, monitoring coverage is highly unequal in global villages.

High-income countries (HIC) typically have dense ground-based monitoring stations that provide broader spatio-temporal coverage. Low- and middle-income countries (LIMCs), particularly in Sub-Saharan Africa (SSA), remain severely under-monitored. According to the WHO ambient air pollution database of 2024, SSA accounts for less than 1% of global monitoring stations (203/40,222) (15). This reflects constraints, including limited policy enforcement, high setting up and operational costs, and shortages of the expertise required for sustained operation of a ground monitoring network (16). This results in limited spatiotemporal coverage of air pollution data, limiting exposure characterization for health assessment, and weakening the design of effective interventions and mitigation strategies (17). This persistent data gap has accelerated interest in alternative air pollution estimation methods, including proxy indicators for air pollution (18).

Proxy indicators are indirect measures used to infer air pollution/quality, including PM_2.5_ concentration, in regions without direct air quality measurements (19). Such indicators leverage open geospatial datasets, such as road network density, visibility, meteorological variables, and land-use/land-cover patterns, to characterize air pollution and infer pollutant concentrations (20). Road network-related proxies are of particular interest as traffic and associated resuspension are important contributors to urban PM_2.5_, and because road data are widely accessible (17). Additionally, literature suggests that integrating traffic type, volume, and distance to major routes using a linear regression model strongly predicts PM_2.5_ concentrations (21). However, while these proxies are widely applied in HIC, their validity in SSA remains insufficiently evaluated. SSA exhibits heterogeneous sources of PM_2.5;_ which cannot be quantified and assessed based on existing sparse ground monitoring systems. Robust validation of road and traffic related PM_2.5_ proxy indicators is therefore necessary to determine whether they can reliably support pollution exposure characterization in SSA.

In this study, we evaluated the validity of three road-network-based proxies for PM_2.5_ exposure: (i) population-weighted Road Network Density (WRND), (ii) Euclidean distance to Highway (EH), and (iii) Euclidean distance from Main roads (EM). The main aim of this study was to evaluate how these proxies performed against measured outdoor-filtered PM_2.5_ personal exposure data from The Gambia, Mozambique, and Kenya, collected as part of the PREgnancy Care Integrating Translational Science, Everywhere PRECISE- DYAD project (22,23).

## 2 Methods

### 2.1 Overall framework

We developed a framework to validate road-traffic proxy indicators using high-resolution personal PM_2.5_ exposure data (Fig. 1). The framework integrates personal PM_2.5_ measurements from the PREgnancy Care Integrating Translational Science Everywhere (PRECISE-DYAD) project, road network data from OpenStreetMap, and gridded population data from WorldPop. All analyses in the study followed the Characterizing Effects of Air Quality In Maternal, Newborn and Child Health (CHEAQI-MNCH) protocol(24), and the use of PRECISE DYAD and PRECISE-HOME data complied with project ethical clearance and guidance.

**Figure 1.**
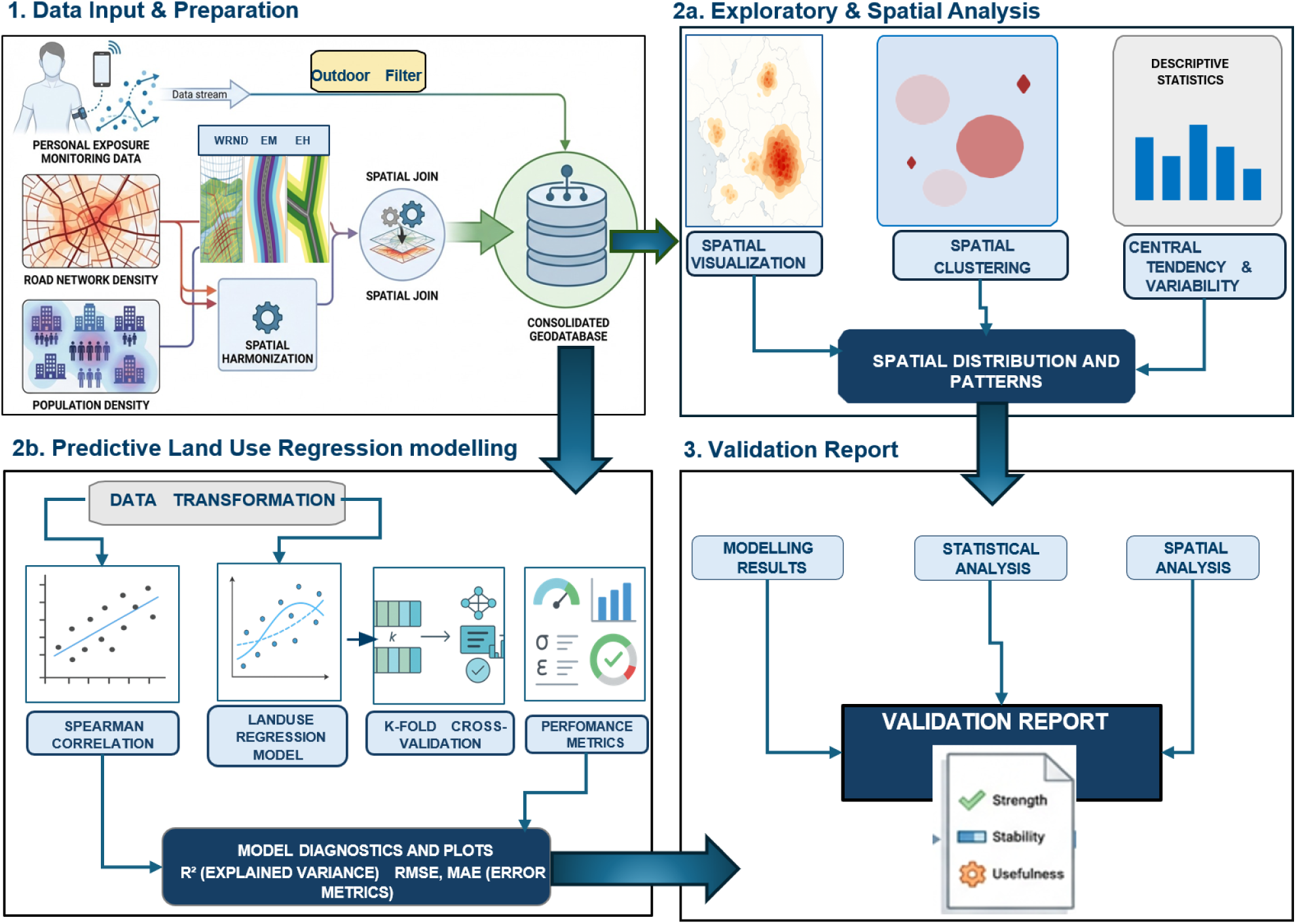
Flow chart of the validation framework developed in the study to validate the proxy indicators in Kenya, The Gambia, and Mozambique.

While the PRECISE-DYAD personal monitoring protocol captures integrated exposure across indoor and outdoor micro-environments, this analysis restricted observations to outdoor movement periods to better isolate the traffic-related component relevant to proxy evaluation.By evaluating these proxies against outdoor PM_2.5_ exposure measurements across multiple SSA settings, we provide a conceptual framework for PM_2.5_ estimation in regions with sparse conventional air quality networks.

### 2.2 Study Sites

The study was conducted in selected villages in three African countries: southern Mozambique, coastal Kenya, and the North Bank region of The Gambia (25,26) (Fig 2). These sites represent contrasting SSA geographies, road connectivity, settlement form, and land use contexts that influence spatial patterns of PM_2.5_ exposure (27,28). In Mozambique, the study villages are located within Manhica District and Xinavane locality, both in Maputo province, an area characterized by intensive large-scale sugarcane agricultural activity (29). In Kenya, the study sites fall within the coastal corridor around Mariakani, and the rural Rabai Sub-County located along the northern corridor, where heavy traffic volumes are a major potential source of PM_2.5_ (30). The Gambia, the study sites are situated in and around Farafenni and within surrounding rural districts, including Illiasa and Sabach Sanjal. These districts are characterized by dense population and a high level of informal transport (31). In general, ground air quality monitoring is limited in these countries and concentrated in a small number of locations, primarily in capital cities (Mozambique: 1 in Boane, Kenya: 1 in Nairobi and 1 in Mombasa, The Gambia: 1 in Banjul).

**Figure 2:**
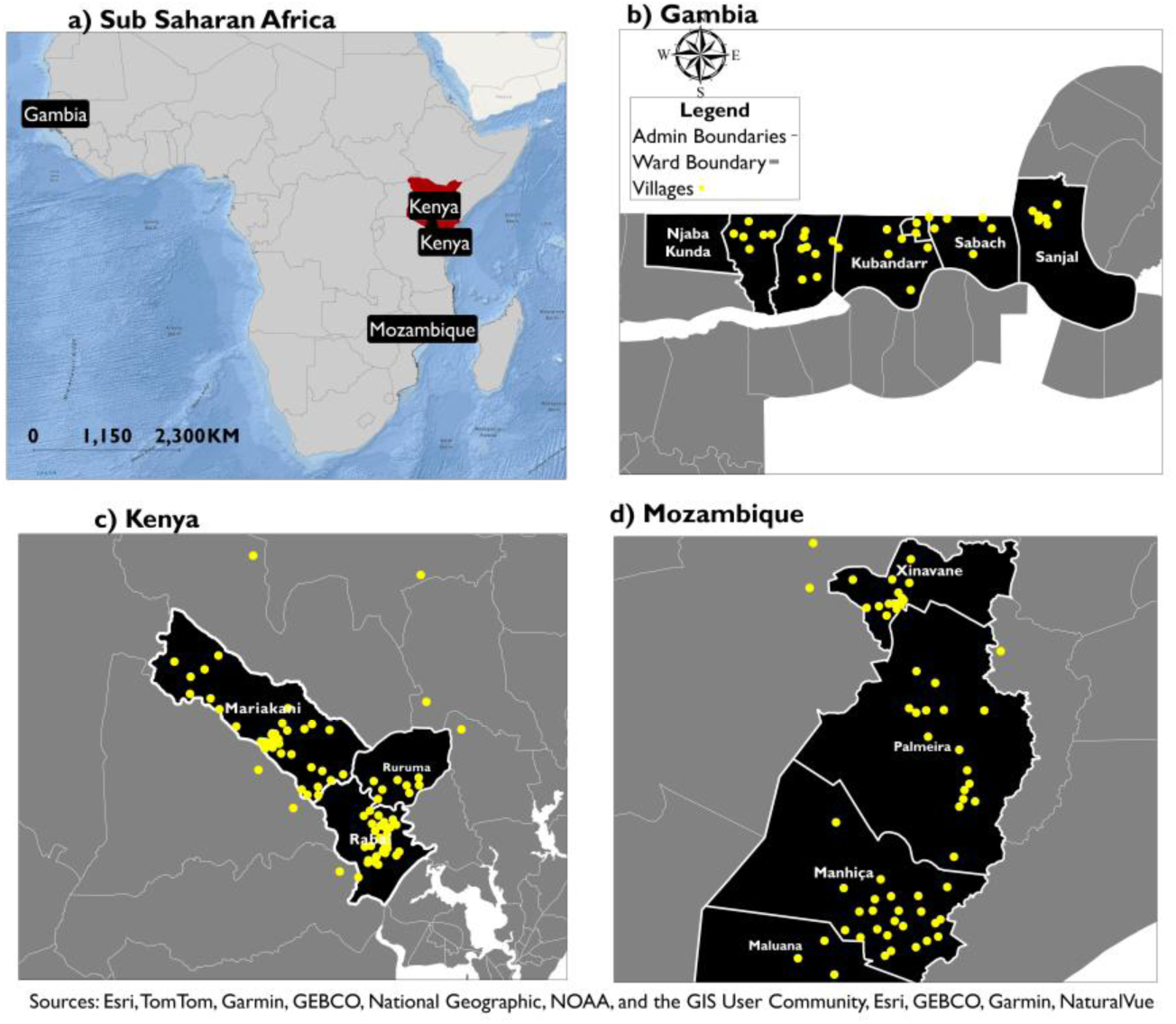
PRECISE study villages located in Kenya, Mozambique, and The Gambia, representing East, West, and Southern Africa regions. Data used to validate proxy indicators was collected in these villages.

### 2.3 Data: Personal exposure and proxies

This study used a secondary high-resolution personal PM_2.5_ exposure dataset from the PRECISE-DYAD project comprising 1,048,576 one-minute observations collected over 12 months (2022-2023). In total, 343 women were recruited in PRECISE-DYAD to carry a personal exposure monitor bag that recorded PM_2.5_ concentrations (µg/m^3^), alongside relative humidity, and air temperature, capturing both indoor and outdoor microenvironments. However, this study focused on outdoor exposure as it better reflects road traffic-related pollution.

To identify outdoor mobility periods and reduce contamination from stationary and indoor time, we applied a spatial displacement threshold of 100 m/min (1.55 m/s). This threshold exceeds the commonly used static-cluster threshold (∼0.83 m/s) used in other studies (32,33), thereby reducing misclassification of non-outdoor periods as outdoor. The selected value was consistent with typical adult walking speed (1.0–1.8 m/s) (34,35), supporting the use of 1.5 m/s to identify sustained outdoor movement.

To improve the robustness of this classification, a spatial validation step was implemented using footprint polygons from OpenStreetMap. Building geometries were retrieved through the Overpass API and used to perform point-in-polygon analysis to determine whether monitoring points occurred inside or outside building footprints. Speed-based classifications were compared with building-based labels using confusion matrix analysis, and discrepancies were corrected using the building-derived classification as ground truth to minimize misclassification of indoor and outdoor environments. After applying these filtering and validation procedures, the recorded observations were retained for subsequent exposure analysis.

We derived a population-weighted road network density (WRND) indicator by integrating road density with population distribution. The inputs included OpenStreetMap road network data (May 2024), village boundary polygons, village centroid point locations, and WorldPop (2018) gridded population data at 100m spatial resolution. We projected the road-network dataset to WGS84 UTM coordinate system to support spatial distance and area calculations. We used ArcMap 10.8 to compute WRND at the neighborhood level by first clipping the road network to community boundaries and generating a regular fishnet grid aligned to the 100m population raster. The total road length within each grid cell was summarized using the cell ID as the aggregation unit, and road network density (RND) was calculated per unit area (km/km^2^). Population counts from WorldPop were extracted and assigned to the corresponding grid cell, producing a weighted Road Network Density score. The wRND score was computed using Eq. 1.

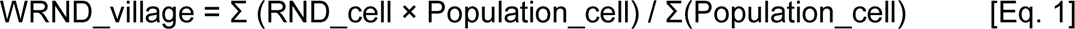

We also derived distance proxies in ArcMap 10.8 by calculating Euclidean (36) (straight-line) distances from village centroids to the nearest main roads (EM) and highways (EH) based on the OpenStreetMap road network. The road network was reprojected to WGS84 UTM Zones to ensure accurate metric distance estimation, and roads were classified by hierarchy to isolate main roads and highways for separate analysis. The Near tool was used to calculate the shortest straight-line distance from each village centroid to the closest road segment, identifying the nearest location along the road geometry.

### 2.4 Data Analysis: Spatial Association and Predictive Modelling of Personal PM2.5 Exposure Using Proxy Indicators

We mapped the spatial distribution of the proxy-PM_2.5_ relationship using proportional symbols in ArcMap 10.8. For each study region, a 5 km hexagonal tessellation grid (37) was generated, and village locations were spatially joined to the tessellation to aggregate village-level attributes within each hexagon. Within each hexagon, PM_2.5_ and proxy indicators (WRND, EM, and EH) were summarized using the mean, producing an aggregated surface of village clusters rather than individual village points. Proxies were then visualized as proportional symbol sizes, while PM_2.5_ was represented using a sequential color gradient (µg/m³), enabling comparison of PM_2.5_ intensity across proxy ranges within and between study regions. Proxy-PM_2.5_ associations were evaluated using Spearman’s rank correlation (38).

We modeled PM_2.5_ as a function of road-traffic proxies WRND, EH, and EM, fitting them individually and jointly within a Land Use Regression (LUR) model Framework. LUR is widely used to estimate spatial variations in pollutant concentrations using proxy geospatial predictors (39,40). LUR was implemented using a random forest (RF) regressor and an ensemble method that combines multiple decision-tree models on bootstrapped samples and averages predictions to reduce variance and overfitting (41). At each node split within a tree, RF considers a random subset of predictors (*mtry*), which decorrelates between trees and improves robustness.

Country-specific RF models were developed for the study countries (Fig. 3). We set the number of trees (ntree) to 50 for computational efficiency and *mtry* to the square root of the number of predictors. The model performance was evaluated using 3 cross-validation folds and summarized by the coefficient of determination (R^2^), root mean squared error (RMSE), mean absolute error (MAE), and mean bias error (13,42,43). Feature importance was computed for the combined models.

**Figure 3:**
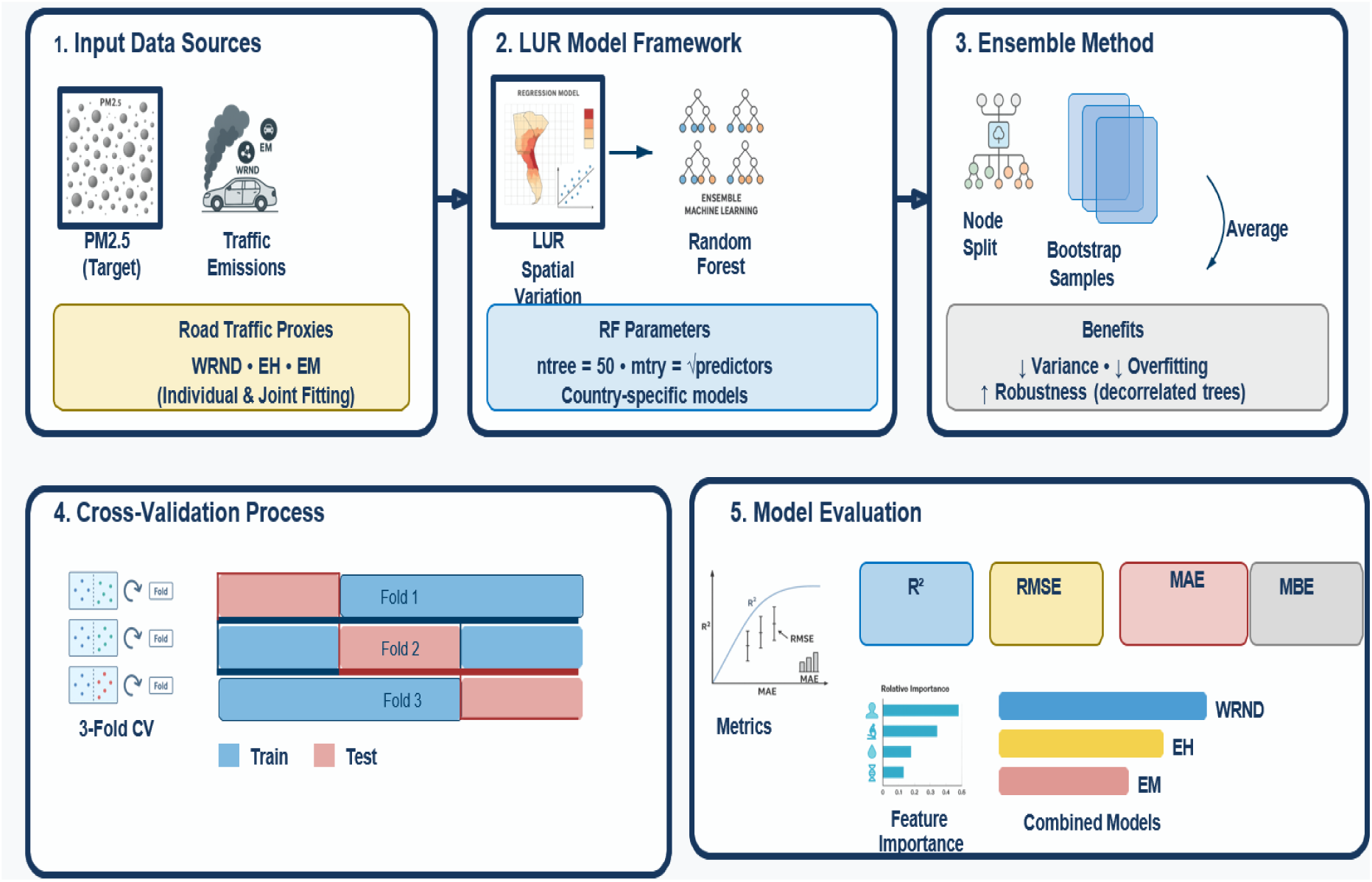
Diagrammatic representation of the random forest framework used to estimate PM_2.5_ across the study sites. For each country, four models were fitted (WRND, EH, EM, and Combined), each with 50 trees. Model outputs for PM_2.5_ were averaged across trees within each model and evaluated separately.

## 3 Results

### 3.1 Proxies and Personal PM2.5 exposure – Summary statistics and spatial distribution

Kenya recorded the highest mean WRND (74.8), alongside PM_2.5_ mean = 31.3 µg/m³ (max = 173.1 µg/m³), EM mean = 0.7 km, and EH mean = 1.3 km (max = 42.5 km) - (Table 1). Mozambique showed the highest mean EH (4.0 km), with PM_2.5_ mean = 19.6 µg/m³ (max = 60.7 µg/m³), WRND mean = 16.1, and EM mean = 1.1 km. The Gambia displayed intermediate proxy levels (WRND mean = 26.1; EM mean = 1.5 km; EH mean = 1.8 km) but the highest PM_2.5_ burden overall (PM_2.5_ mean = 49.4 µg/m³; max = 220.7 µg/m³)

**Table 1:** Summary statistics PM*_2._*_5_ (µg/m³), WRND, EM(km), and EH(km) across study sites over the full analysis period.

| Variable | Country | Mean | Std | Min | 25% | Median | 75% | Max |
| --- | --- | --- | --- | --- | --- | --- | --- | --- |
| PM<sub>2.5</sub> | Gambia | 49.4 | 42.0 | 9.0 | 24.5 | 35.5 | 66.5 | 220.7 |
|  | Kenya | 31.3 | 35.7 | 4.8 | 9.6 | 16.3 | 34.1 | 173.1 |
|  | Mozambique | 19.6 | 11.8 | 2.3 | 10.4 | 18.2 | 24.1 | 60.7 |
| WRND | Gambia | 26.1 | 25.1 | 5.9 | 17.0 | 20.7 | 25.7 | 152.5 |
|  | Kenya | 74.8 | 46.8 | 0.3 | 41.5 | 75.0 | 98.2 | 198.0 |
|  | Mozambique | 16.1 | 7.6 | 2.5 | 8.8 | 17.4 | 21.2 | 36.5 |
| EM(km) | Gambia | 1.5 | 1.6 | 0.2 | 0.4 | 0.7 | 2.1 | 6.7 |
|  | Kenya | 0.7 | 0.9 | 0.0 | 0.1 | 0.5 | 0.9 | 7.0 |
|  | Mozambique | 1.1 | 1.0 | 0.0 | 0.5 | 0.7 | 1.5 | 5.4 |
| EH (km) | Gambia | 1.8 | 1.6 | 0.3 | 0.5 | 1.7 | 2.5 | 6.7 |
|  | Kenya | 1.3 | 4.2 | 0.0 | 0.2 | 0.6 | 1.2 | 42.5 |
|  | Mozambique | 4.0 | 4.2 | 0.0 | 0.8 | 1.9 | 7.0 | 17.86 |

The bivariate spatial distribution of PM_2.5_ and WRND across the village study villages aggregated into tessellations. is shown in Figure 4 In The Gambia (Fig. 4a), villages along Njaba Kunda/No Kunda and Farafenni have average to higher PM2.5 concentrations, ranging from low to moderate (0–20). In Kenya (Fig. 4b), villages in Mariakani and Rabai show mixed spatial patterns in which elevated PM_2.5_ occurs across multiple WRND classes. In Mozambique (Fig. 4c), Xinavane exhibits co-located higher values of PM_2.5_ and WRND; these clusters extend through some villages in Palmeria but occur across both lower and higher WRND classes.

**Figure 4:**
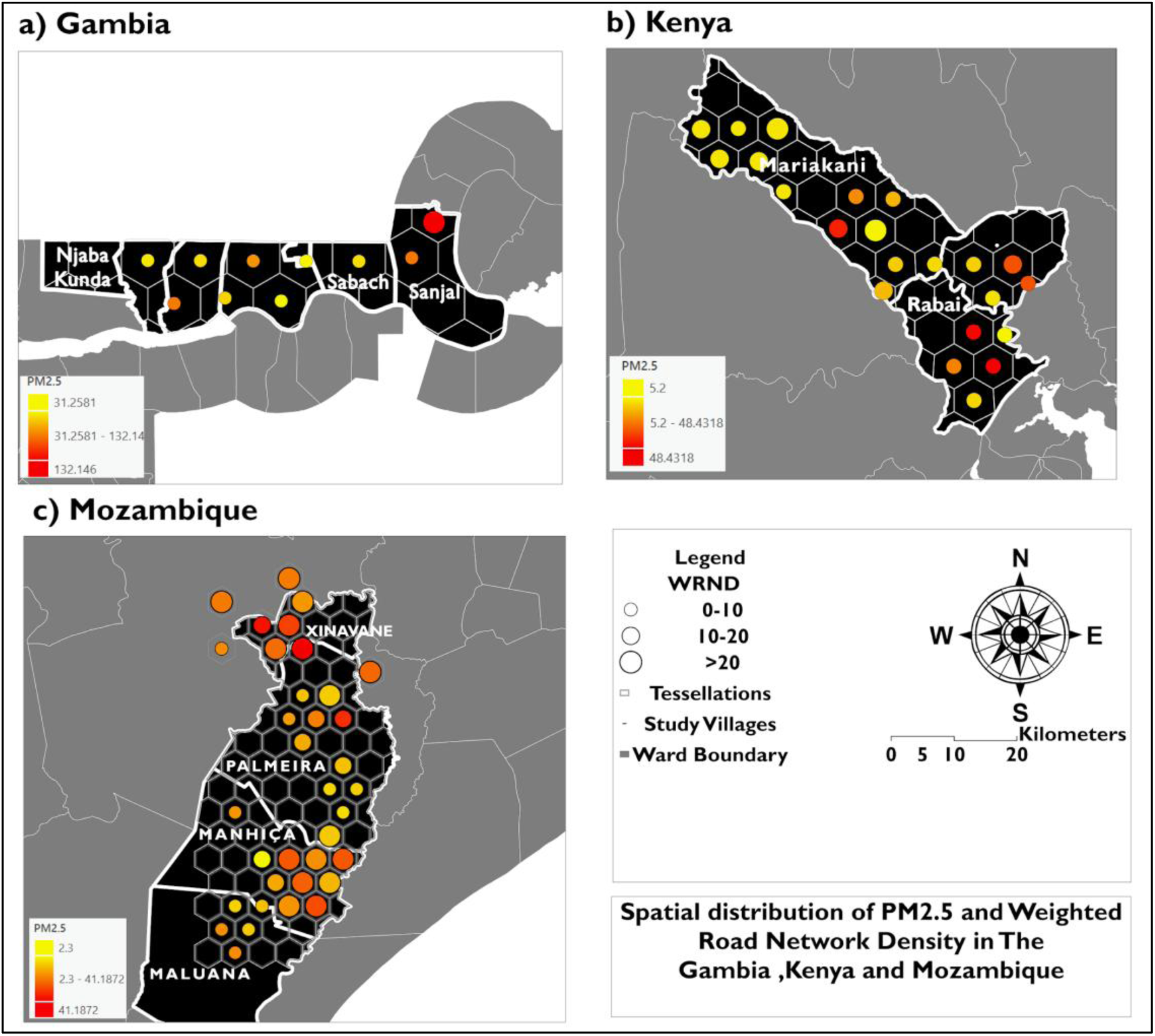
Spatial distribution of aggregated PM_2.5_ and weighted road network density (WRND) across hexagonal tessellations in (a) The Gambia, (b) Kenya, and (c) Mozambique. Hexagons represent aggregation units of the study villages. The yellow-to-orange color gradient represents PM2.5 (µg/m³), and proportional symbols represent WRND classes (0–10, 10–20, and >20) within each hexagon.

the spatial distribution of the PM_2.5_-EM relationship in the study villages. is shown in figure 5 In The Gambia (Fig. 5a), villages in Sabach and Farafenni show higher PM_2.5_ concentrations, including low EM classes, while villages more than 20 km from Sanjal show elevated PM_2.5_ at greater distances. In Kenya (Fig. 5b), Mariakani and Rabai exhibit mixed patterns in which higher PM_2.5_ occurs in both low EM (0–2 km) and intermediate EM (2–4 km) hexagons. The results highlight that in Mozambique (Fig. 5c), villages in Xinavane exhibit higher PM2.5 concentrations across multiple EM classes, including some hexagons with larger EM (>3 km), contradicting a distance-decay effect in PM2.5 with increasing EM. Overall, across the study villages, the PM_2.5_–EM relationship is heterogeneous across and within sites, consistent with context-dependent proxy performance.

**Figure 5:**
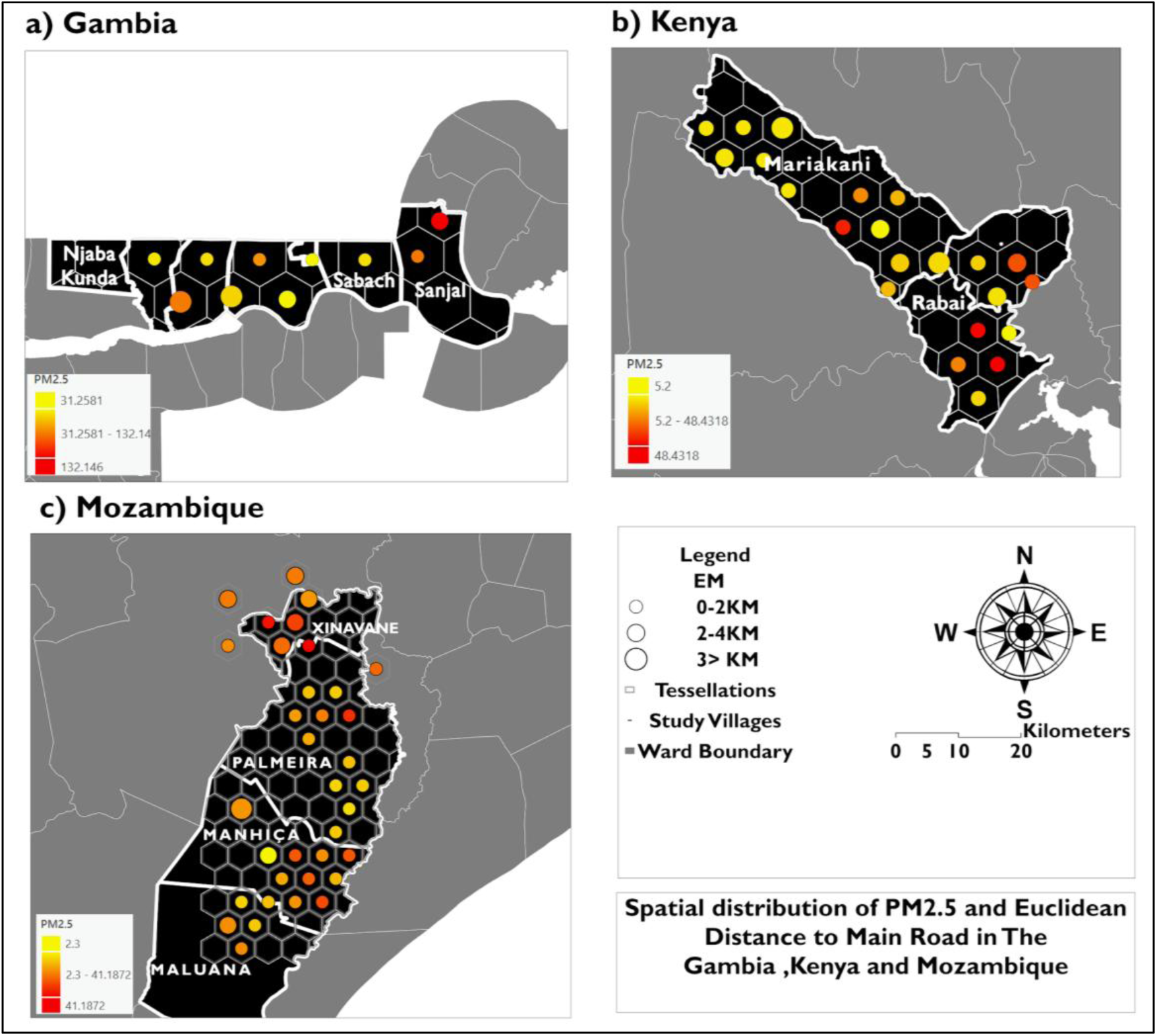
Spatial distribution of aggregated PM_2.5_ and Euclidean distance to main roads (EM) across hexagonal tessellations in (a) The Gambia, (b) Kenya, and (c) Mozambique. Hexagons represent aggregation units of the study villages. The yellow-to-orange color gradient represents mean PM_2.5_ (µg/m³), and proportional symbols represent mean EM classes (0–2 km, 2–4 km, and >3 km) within each hexagon.

The spatial distribution of the PM_2.5_–EH relationship in the study villages, is shown in figure 6 as hexagon tessellations. In The Gambia corridor villages in Njaba Kunda and Sabach show moderately elevated PM_2.5_ at low EH, aligning more closely with the proximity-to-highways assumption. In Kenya (Fig. 6b), Mariakani shows a proximity effect, with aggregated low-EH values corresponding to higher PM_2.5_ concentrations. In contrast, village Rabai shows higher PM_2.5_ at higher EH. In Mozambique (Fig. 6c), Xinavane shows a direct positive relationship between higher PM_2.5_ and greater highway distances, contradicting the expected decline in PM_2.5_ with increasing EH.

**Figure 6:**
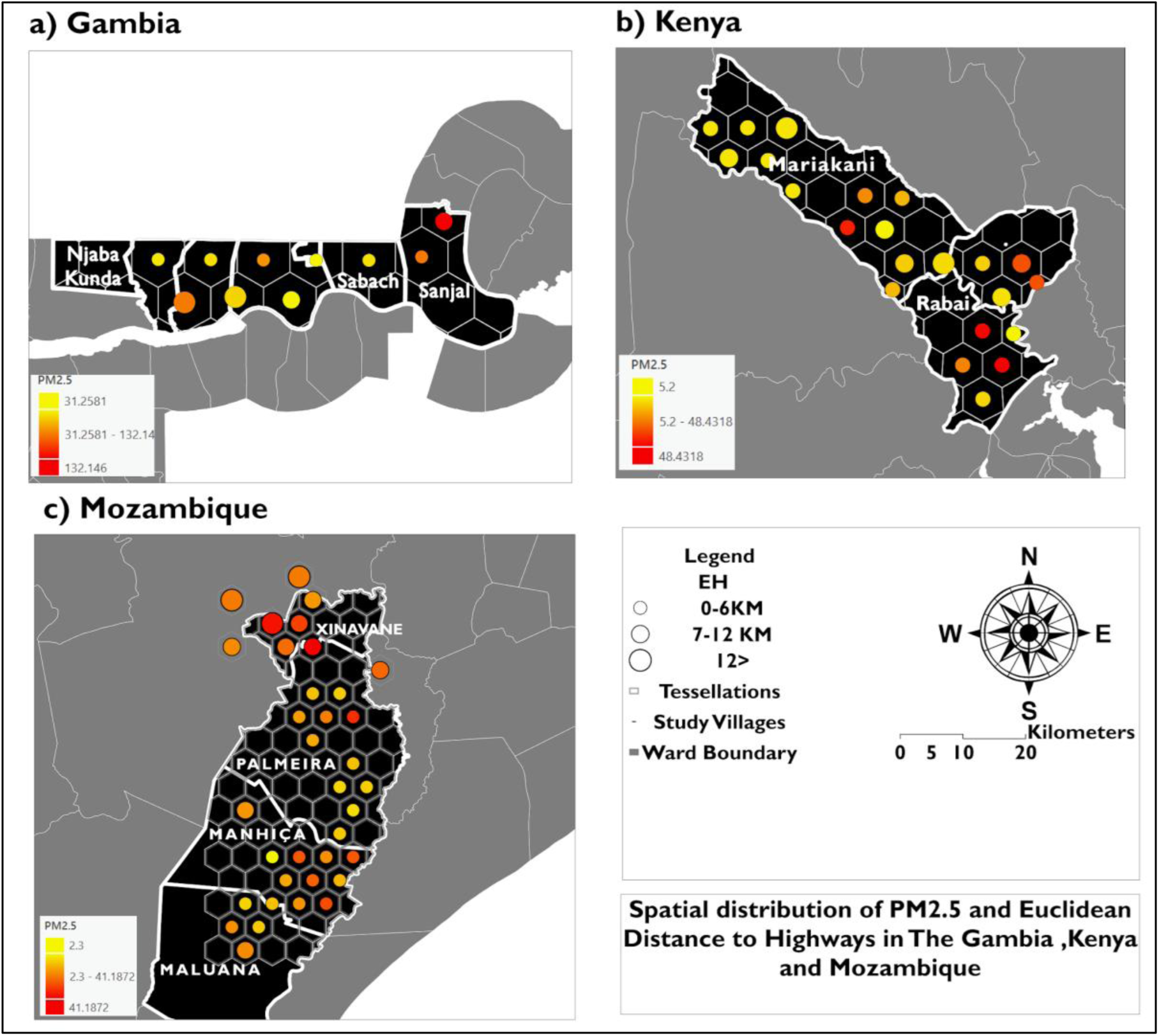
Spatial distribution of aggregated PM_2.5_ and Euclidean distance to highways (EH) across hexagonal tessellations in (a) The Gambia, (b) Kenya, and (c) Mozambique. Hexagons represent the aggregation units. The yellow to orange color gradient represents mean PM_2.5_ (µg/m³), and proportional symbols represent mean EH (km) within each hexagon.

Proxy-PM_2.5_ associations in Figure 7 indicate limited and context-specific statistical relationships. WRND was positively and significantly associated with PM_2.5_ in Mozambique (r = 0.351; *p = 0.005*), but associations were weak and non-significant in Kenya (r = −0.041; *p = 0.673*) and The Gambia (r = −0.020; *p = 0.909*); the pooled (“global”) association was near zero (r = 0.032; *p = 0.648*). EM showed weak, non-significant associations across all settings (Kenya r = −0.103; *p = 0.280*; The Gambia r = 0.146; *p = 0.410*; Mozambique r = −0.038; *p = 0.766*), with an overall weak negative Global association ( r = −0.008; *p = 0.912*). EH exhibited contrasting behavior, with a significant negative association in Kenya (r = −0.224; *p = 0.018*) and a positive association in Mozambique (r = 0.292; *p = 0.020);* whilst association in The Gambia had positive but marginal (r = 0.335; *p = 0.053*).

**Figure 7:**
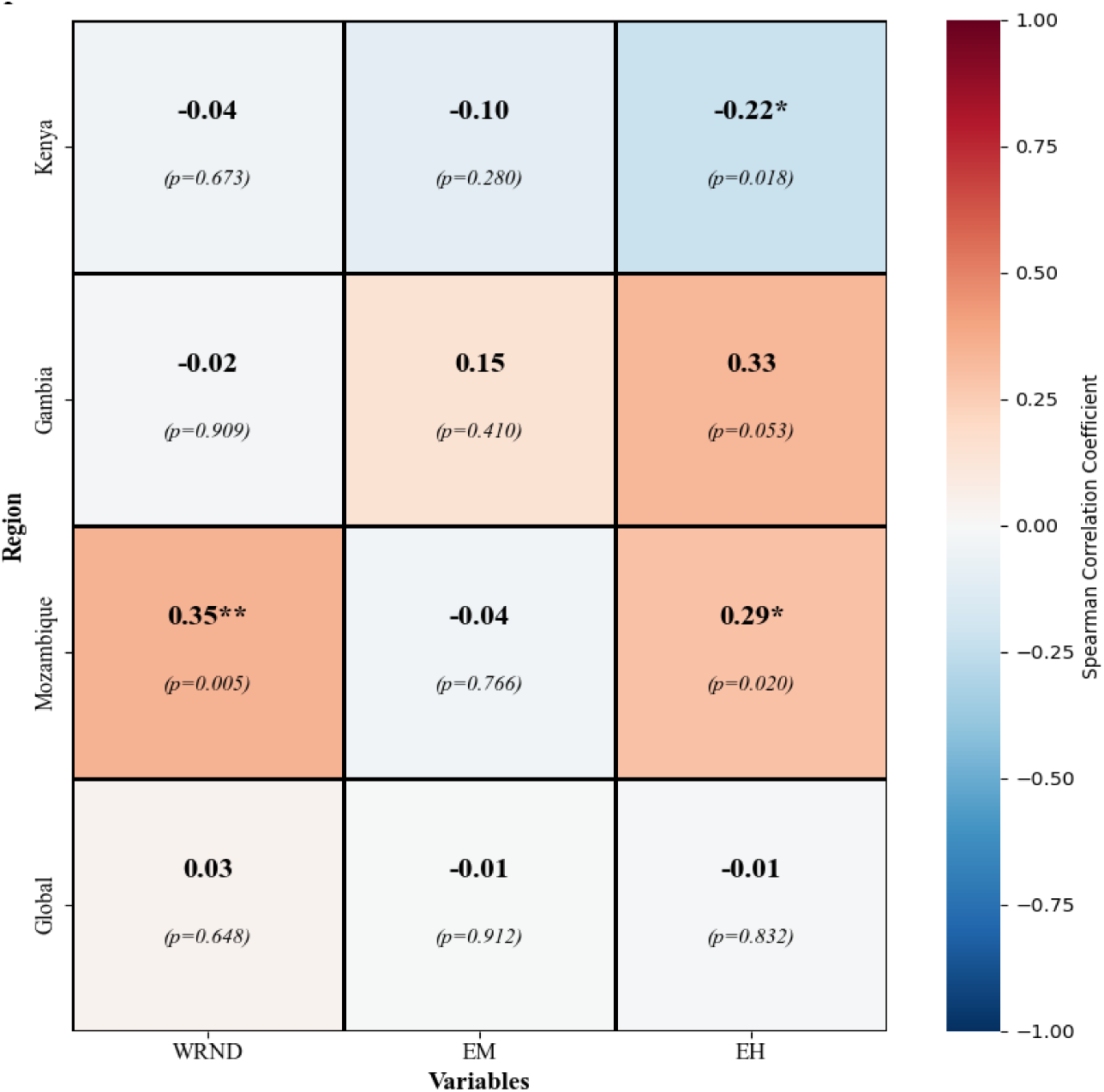
Spearman correlations between proxy indicators (WRND, EH EM) and pollutants (PM_2.5_) in Mozambique, Kenya, and the Gambia. The shades of blue indicate negative correlations, while those of red indicate positive correlations. Significance is denoted by asterisks: * p < 0.05, ** p < 0.01, *** p < 0.001.

### 3.2 Regression analysis of Personal PM2.5 exposure and proxy indicators

The observed versus predicted values for the RF models, using single and combined models across the study sites is shown in figure 8. The performance metrics of the models are in Table S2 and Figure S1.

**Figure 8:**
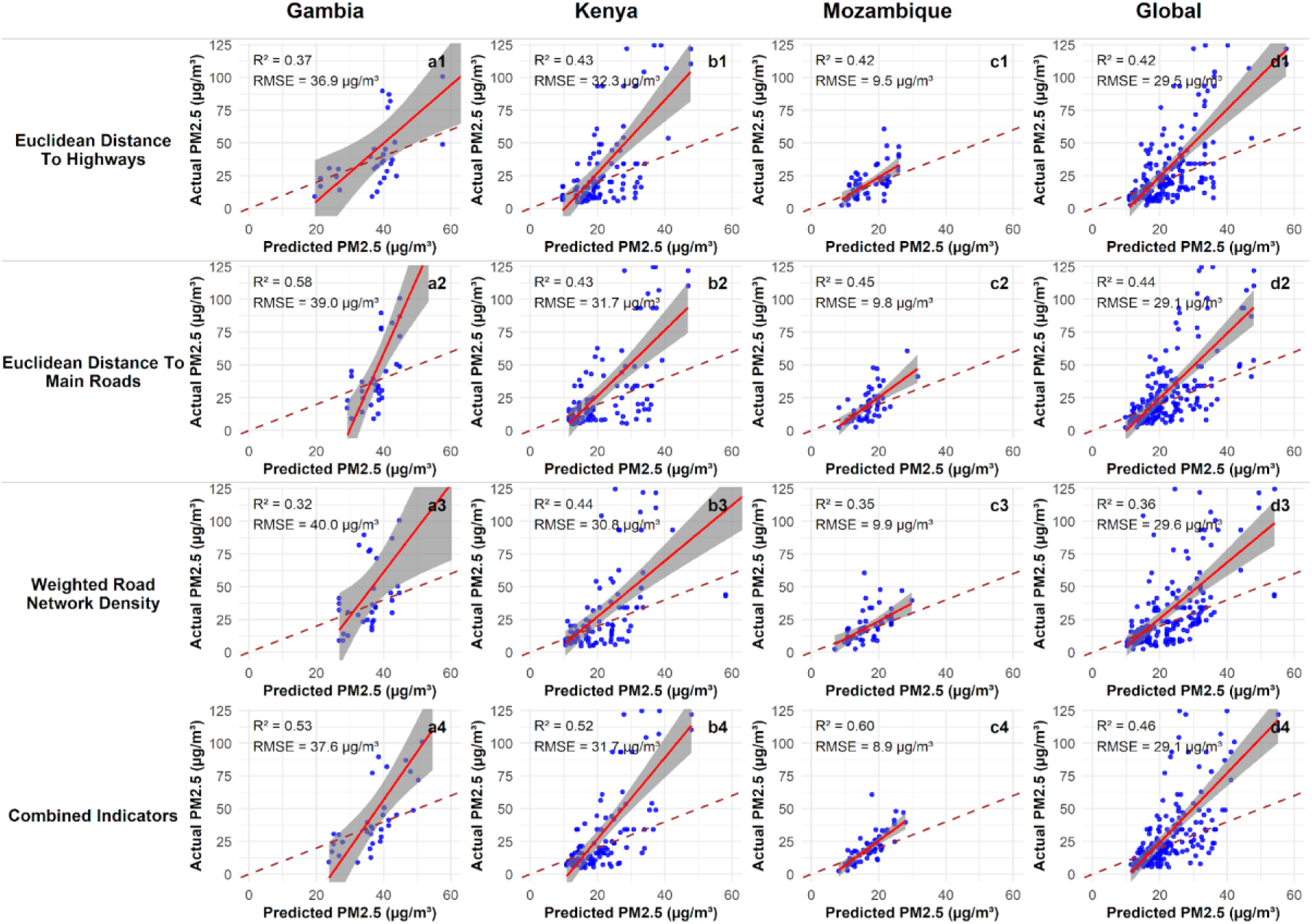
Observed versus predicted PM_2.5_ from Random Forest models using single and combined proxy indicators across study countries and the whole dataset. The panels show the PM_2.5_ model performance for each country, annotated with R^2^ and RMSE.

EM only models’ regression ranged from The Gambia (R^2^=0.37 ; RMSE = 36.97 µg/m^3^), Kenya (R^2^ = 0.43; RMSE = 32.3 µg/m^3^), and Mozambique (R^2^ = 0.42; RMSE= 9.5µg/m^3^) and Global performing moderately as well (R^2^=0.42, RMSE= 32.3µg/m^3^). EH-PM_2.5_ models were the strongest single proxy in performance (R^2^= 0.58; RMSE = 39.0 µg/m^3^) and remained comparable in Kenya (R^2^ = 0.43; RMSE = 31.7 µg/m^3^) and Mozambique (R^2^ = 0.45; RMSE = 9.8 µg/m^3^), with Global R^2^ = 0.44 (RMSE = 29.1 µg/m^3^).

WRND-only models showed weaker performance in The Gambia (R^2^ = 0.32; RMSE = 40.0 µg/m^3^) but were like EM/EH in Kenya (R^2^= 0.44; RMSE = 30.8 µg/m^3^) and moderate in Mozambique (R^2^= 0.35; RMSE = 9.9 µg/m^3^), with Global R^2^ = 0.36 (RMSE = 29.6 µg/m^3^). Combining indicators improved performance in Kenya (R² = 0.52; RMSE = 31.7 µg/m³) and Mozambique (R^2^ = 0.60; RMSE = 8.9 µg/m^3^) and increased pooled performance (Global R^2^= 0.46; RMSE = 29.1 µg/m^3^), although in The Gambia the combined model (R^2^ = 0.53; RMSE = 37.6 µg/m^3^) did not exceed the EM-only model.

Mean bias Error (MBE) plots for PM_2.5_ are shown in Fig.9.Combined proxy models showed the smallest and most stable biases across countries in Kenya; the combined biases ranged from mean ±2–5µg/m^3^ for low and mid-range(0-40µg/m^3^) concentration bins. Whilst in The Gambia and Mozambique, MBE exceeded 40 µg/m³ in the combined model at higher PM2.5 concentrations; the deviations were +20 µg/m³ at higher-range concentrations.

**Figure 9:**
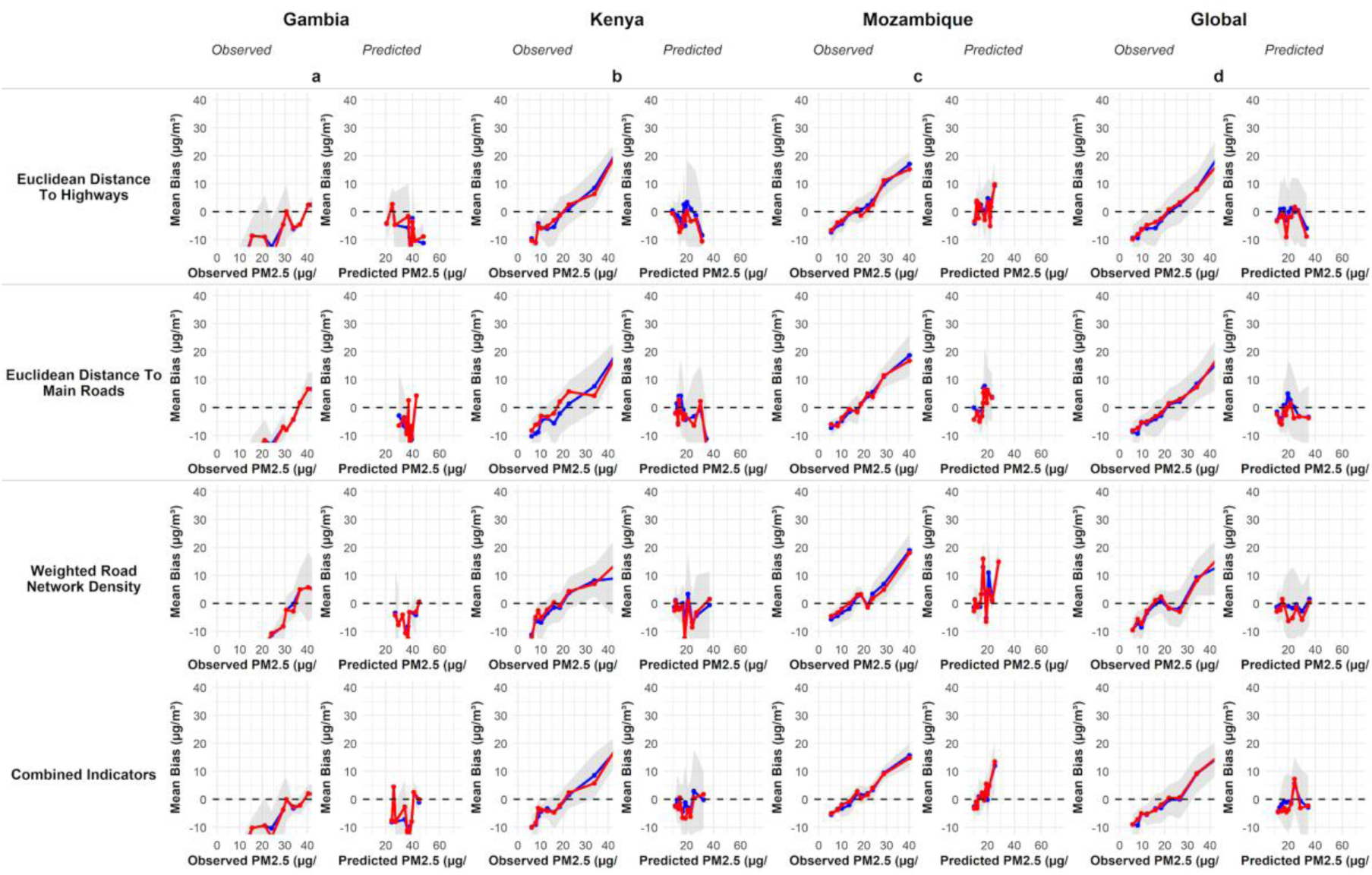
Mean bias error (MBE) plots for random forest PM_2.5_ predictions across study countries and the global dataset. The black line shows mean bias, the red line shows median bias, and the grey shading indicates the 95% confidence interval for mean bias. The plots show that combined proxy models exhibit the lowest overall bias, though bias increases to +10 µg/m³ at the highest concentrations.

## 4 Discussion

In this study, we evaluated the validity of road traffic-related proxy indicators in estimating PM_2.5_ pollution exposure across three SSA countries (The Gambia, Kenya, and Mozambique), characterized by limited monitoring infrastructure. Beyond validating the study site-specific validation, our analysis provides a structured framework for evaluating the utility of proxy indicators in representing PM_2.5_ in areas with limited monitoring infrastructure.

### 4.2 Spatial distribution of proxy indicators and Personal PM2.5 exposure

In The Gambia, areas characterized by higher WRND and low EM exhibited higher PM_2.5_ concentrations, consistent with evidence linking dense road networks with elevated PM_2.5_ levels in urban and peri-urban environments (44). Similar spatial relationships between proximity to major roads and PM_2.5_ have been demonstrated in previous proximity-based analyses in Ireland (45).

Additionally, Traffic dispersion modelling in road-dense corridors yielded the same results, supporting the use of road proximity as a proxy of PM_2.5_ exposure (46). In contrast, Mozambique’s PM_2.5_ concentrations were less dependent on EM. They remained elevated even greater distances from main roads, highlighting that local sources were dominant and emanated from non-traffic processes. This finding mirrors findings from the same Xinavane region, where sugarcane burning is a major contributor to PM_2.5_ (47). In Kenya, higher PM_2.5_ levels from villages farther from main roads also suggest on-road traffic sources, as identified in other studies, including biomass burning and industrial activities (48).

### 4.3 Statistical Relationship of proxy indicators and Personal PM2.5 exposure

Overall, WRND showed moderate alignment with PM_2.5,_ in line with studies reporting positive road-PM_2.5_ associations in urban areas (49,50). However, weaker WRND-PM_2.5_ correlations in Kenya and The Gambia indicate that WRND alone is insufficient in some settings, suggesting stronger roles for land use, meteorology, and non-traffic emissions in shaping PM_2.5_ variability (51).

Notably, the positive correlation between PM_2.5_ and EH in The Gambia and Mozambique contradicts the typical distance-decay pattern observed in many LMICs (52). However, this reflects confounding non-traffic sources (biomass and agricultural burning) that elevate PM_2.5_ away from major highways. Overall, road proxies can inform PM_2.5_, but validity is context-dependent and improved by adding environmental covariates.

### 4.4 The regression analysis of Proxy indicators in predicting Personal PM2.5 exposure

Single proxy RF models showed limited predictive power for PM_2.5_ across all study sites, reflecting exposure variability unlikely to be captured by a single road-based variable. (53,54). Further, this finding aligns with studies in Italy, which also found that single proxy models had limited predictive capacity for PM_2.5._ (50). In particular, reliance on proximity-only measures (e.g., EM) demonstrated limited robustness, reflecting sensitivity to local spatial heterogeneity and shifting emission sources that complicate exposure modelling in urban and peri-urban environments (55).

In contrast, combining proxy indicators significantly improved PM_2.5_ prediction, highlighting the value of integrating spatial indicators, such as road network density and proximity to major roads, to represent exposure variability. This finding aligns with work from Cape Town, South Africa, and other LMIC regions, showing that multi-indicator models improve PM_2.5_ predictions relative to isolated proxies (56–58). Importantly, the pooled (“global”) analysis indicated moderate overall skill and further improvement when proxies were combined, suggesting that aggregating data across settings can stabilize estimation and capture shared proxy–PM_2.5_ structure that is not consistently recoverable within individual sites.

Nevertheless, predictive performance varied across countries, with weaker results in Kenya suggesting that road traffic proxies alone may not adequately capture PM_2.5_ spatio-temporal variability. This highlights the models’ limitations and the need for localized calibration and greater integration of environmental factors.

The feature of importance results further indicated that the dominant proxy contribution differs across countries. This reinforced the idea that proxy validity is context-specific and that modelling strategies should be tailored to local sources, mixtures, and spatial structures (59,60). Consistent with this are previous studies in Thailand that demonstrated that integrating satellite, environmental variables, and traffic data improves ground-level PM_2.5_ estimates, supporting a combined-data approach for exposure assessment (17).

### 4.5 Implications of Validation

These findings underscore the importance of validating proxy indicators of PM_2.5_ exposure. While some proxies demonstrated utility, inconsistent associations across countries indicate that single indicators are insufficient, and a multi-proxy approach is more robust. Prior evidence shows that incorporating meteorological variables, land-use patterns, and satellite-derived aerosol optical depth (AOD) data can improve estimates of PM_2.5_ exposure from diverse pollution sources (61,62).

Moreover, our study demonstrates the effectiveness of RF-based LUR models for estimating PM_2.5_ exposure in resource-constrained settings. However, between-country variability highlights the need for local calibration and expanding predictor sets where road proxies alone incompletely represent exposure dynamics.

## 5 Conclusion and Recommendations

This validation framework integrated personal exposure measurements with open geospatial data to assess the robustness of road-traffic proxies for PM_2.5_ in settings with limited monitoring. Overall, WRND proved to be the most consistent strong predictor of PM_2.5_. However, positive associations between EH and PM_2.5_ in The Gambia suggested significant contributions from non-traffic sources such as marketplaces. Underscoring the limits of pure road-based proxies. Spatial patterns also indicated elevated PM_2.5_ concentration both near and far from main and highway roads, highlighting the need for multi-source exposure models.

The LUR RF model results show that combining road proxies improved PM_2.5_ exposure estimates, but performance is likely to improve further through the integration of meteorological and remote-sensing covariates.

### Based on these findings, we recommend

The development of hybrid models that combine LUR with satellite and meteorological datasets to improve PM_2.5_ exposure estimation in rapidly urbanizing regions of Sub-Saharan Africa. We also recommend expanding air quality personal and global monitoring sensor networks beyond urban centers to capture non-traffic PM_2.5_ pollution sources that would support calibration and validation. From this study, we have seen the need to integrate road traffic exposure models into urban planning to reduce vehicular emissions while promoting clean energy solutions. There is a need to test and scale this framework to extend PM_2.5_ exposure characterization across additional Sub-Saharan African countries.

## Data Availability

1.The road network and population data underlying the proxy indicators construction presented in the study are available from https://www.openstreetmap.org and https://www.worldpop.org/
2.Personal exposure data cannot be shared publicly because it consisting of location of participants. Data are available from the PRECISE Network Institutional Data Access / Ethics Committee contact via https://precisenetwork.org/ for researchers who meet the criteria for access to confidential data.

## 6 Acknowledgements

We acknowledge the contributions of the women participants in Mozambique, Kenya, and The Gambia, as well as the research teams involved in the PRECISE-Home, which was funded by the Spanish Ministry of Science and Innovation grant MCIN/AEI/10.13039/501100011033, and the PRECISE-DYAD funded by the NIHR–Wellcome Partnership for Global Health Research Collaborative Award, reference 217123/Z/19/Z.

**Funding Acknowledgement** Research was supported by the Fogarty International Centre and National Institute of Environmental Health Sciences (NIEHS) and OD/Office of Strategic Coordination (OSC) of the National Institutes of Health under Award Number U01ES036146-0. The content is solely the responsibility of the authors and does not necessarily represent the official views of the National Institutes of Health.

