## Supplementary figures and images for "Performance of Road-Traffic-Based Exposure Proxies Against Personal PM_2.5_ Measurements in Three Sub-Saharan African Countries"

### Figure S1

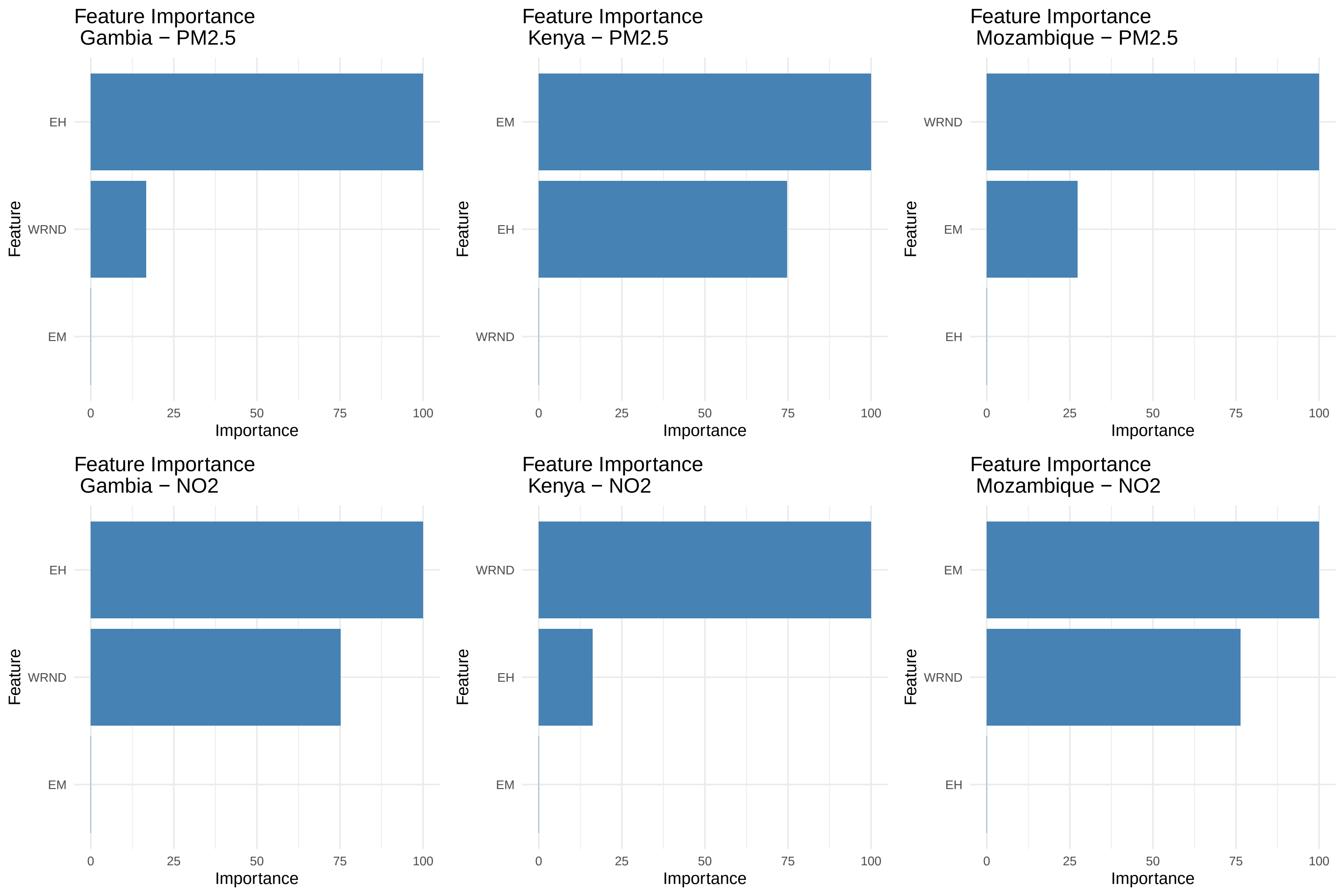
